# Low-dose interleukin-2 for recurrent early pregnancy loss: a proof-of-concept study

**DOI:** 10.1101/2025.06.13.25328550

**Authors:** Arsene Mekinian, Noémie Abisror, Chloé Mcavoy, Claire Ribet, Roberta Lorenzon, Eric Vicaut, Michelle Rosenzwajg, David Klatzmann

## Abstract

**Background:** Regulatory T cells (Tregs) are essential for maternal-fetal tolerance, and their deficiency has been implicated in immune-mediated pregnancy loss. Low-dose interleukin-2 (IL-2_LD_) selectively expands and activates Tregs and has shown efficacy in murine models of spontaneous immune-mediated miscarriage. This proof-of-concept study assessed the safety and efficacy of IL-2_LD_ in women with unexplained recurrent early pregnancy loss (uREPL).

**Methods:** In the FACIL-2 open-label trial (NCT03970954), 15 women with ≥5 prior uREPLs received 3 MIU/day of IL-2 for 5 days per cycle, starting 10 days after menses onset. Up to five cycles were administered if pregnancy was not achieved. Treg frequency was measured by flow cytometry, and clinical outcomes were recorded. An additional 9 patients with similar profiles were treated under compassionate use without immunomonitoring.

**Results:** In FACIL-2 participants, IL-2_LD_ significantly increased circulating Tregs, with a mean 2.0-fold rise at day-8 (p<0.001), primary endpoint reached) and a sustained elevation at day-29. Among the eight pregnancies, four were viable at 12 weeks, resulting in three live births and one late miscarriage at 20 weeks. Successful pregnancies were associated with greater Treg expansion (day-0 to day-8: 2.87 vs. 1.64-fold, p=0.03; day-0 to day-29: 1.71 vs. 1.24-fold, p=0.008). In the compassionate group, five pregnancies yielded two live births. All newborns were healthy. Safety signals were generally mild and manageable.

**Interpretation:** IL-2_LD_ safely expanded Tregs and achieved a 46% rate of viable pregnancies in a high-risk population, and despite the short treatment duration. These results support the further investigation of IL-2_LD_ as an immunotherapy for uREPL in a controlled trial with extended treatment through early gestation.

## INTRODUCTION

Recurrent early pregnancy loss (REPL), defined as the loss of three or more pregnancies before 14 weeks of amenorrhea, affects approximately 1% = of women of reproductive age ^1,2^. While REPL may result from anatomical, genetic, hormonal, infectious, or thrombophilic causes, a substantial proportion of cases remain unexplained after standard evaluation—these are termed unexplained REPL (uREPL). In women with five or more prior losses, the likelihood that miscarriage is due to fetal aneuploidy diminishes significantly—from 63% after two losses to 39% after five. This shift increases the probability of maternal causes, notably immune dysregulation leading to reduced tolerance to the fetus ^1^.

Immune-mediated uREPL is thought to result from an imbalance between effector T cells (Teffs) and regulatory T cells (Tregs), disrupting maternal-fetal tolerance ^3–5^. Multiple studies have reported increased Th1 and Th17 responses, elevated cytotoxic activity, and reduced anti-inflammatory cytokine production in the blood and endometrium of women with REPL^5–8^. In addition, a decrease in Tregs cells in the blood and endometrium and a reduced capacity to secrete IL-10 and TGF-β have been observed in these patients ^3,5,9,10^.

The role of Tregs in implementing maternal-fetal tolerance and preventing immune rejection of the embryo or fetus is further substantiated by the observation that Tregs-depleted mice reject allogeneic fetuses, whereas adoptive transfer of Tregs promotes fetus survival ^11–16^. Furthermore, a rare population of uterine-resident Tregs have been shown to have trophic functions that support fetal growth ^17^. The role of Tregs in pregnancy is also supported by the observation that normal pregnancy is accompanied by an increase in maternal Tregs^18^.

These findings position Treg insufficiency as a plausible therapeutic target in immune-mediated uREPL. Low-dose interleukin-2 (IL-2_LD_) has emerged as a potent and selective activator of Tregs in humans, with proven safety and clinical efficacy in autoimmune diseases ^19–30^. Notably, while high dose IL-2 regimens stimulate Teffs and NK cells, low-dose IL-2 regimens specifically promote Treg expansion and function, without affecting effector responses^31^. In the CBA/J x DBA/2J mouse mating combination, a classical murine model for recurrent spontaneous abortion due to Treg deficiency, IL-2_LD_ has been shown to mitigate infertility and restore the normal pregnancy rate^12,32^.

Building on these findings, we conducted the FACIL-2 open-label proof-of-concept trial to evaluate the immunological and clinical impact of IL-2_LD_ in women with severe uREPL. We hypothesized that peri-implantation IL-2_LD_ would enhance maternal Treg responses and improve pregnancy outcomes in this high-risk population.

## METHODS

### FACIL-2 study design, participants and treatment

FACIL-2 was an open monocentric proof-of-concept trial evaluating the tolerance and the efficacy of IL-2_LD_ in women with REPL. The study was approved by the institutional review board CPP Ile de France III and performed in accordance with the Declaration of Helsinki and good clinical practices. FACIL-2 is registered as “*Low-dose Interleukin-2 in Women With Unexplained Miscarriages*” with ClinicalTrials number NCT03970954.

Patients were recruited from January 2021 to July 2023 and were treated and followed up at the Saint Antoine hospital in Paris. The main inclusion criteria were: women from 18 to 40 years old with at least 5 consecutive recurrent early pregnancy loss before 14 weeks of amenorrhea and unexplained after the usual check-up; volunteer to participate in the trial and having given written consent after appropriate information. The main exclusion criteria were: uterine or pelvic abnormality: uterine malformation, intracavitary fibroid, synechiae, polyp, hydrosalpinx; balanced translocations in both spouses; type I or II diabetes; Sickle cell disease; constitutional or acquired thrombophilia or autoimmunity (protein deficit C, S, ATIII, homozygous factor V or II deficiency, antiphospholipid syndrome, antithyroid antibodies positive, celiac disease, hyperhomocysteinemia); decreased ovarian reserve (anti-mullerian hormone (AMH) <1 ng/ml); antral follicular counts (AFC) < 4 (**table S1**)

Among 18 included patients for the study, 15 were treated with IL-2_LD_ and completed at least one treatment cycle (**Figure 1**). Written informed consent was obtained from all participants before enrolment in the study. Patients received IL-2_LD_ in the form of Aldesleukin (Proleukin® 18 MIU, Novartis Pharma SAS, Rueil-Malmaison, France) provided by the central pharmacy of the Assistance Publique-Hôpitaux de Paris and diluted to the dose of 3MIU/injection (or lower in case of dose reduction) by a pharmacist.

**Figure 1:**
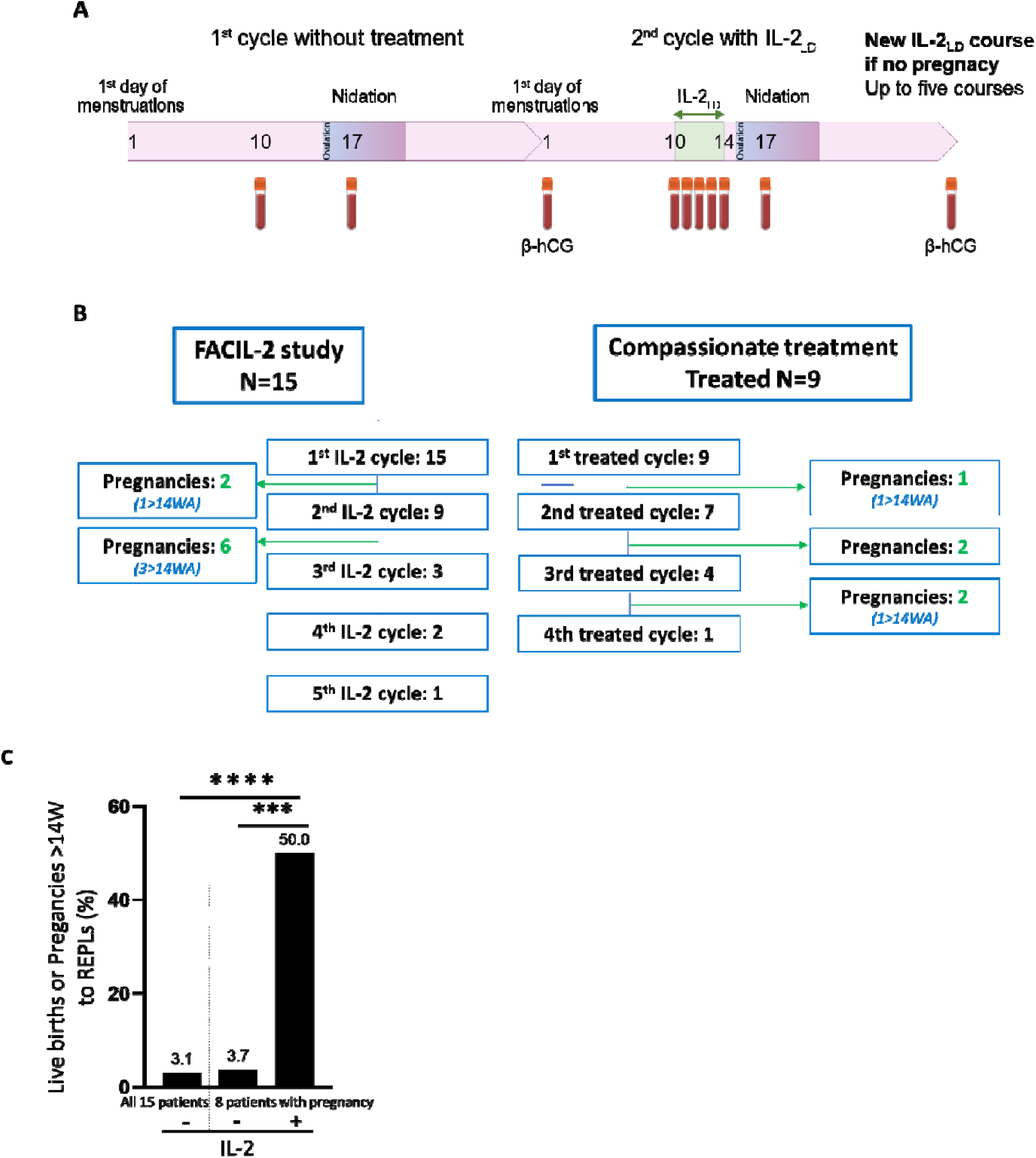
A) Study design; B) Trial profile showing FACIL-2 and compassionate use cohort recruitment and outcomes; C) Ratio of live births or pregnancy >14W to REPLs in all 15 women before inclusion in the FACIL-2 trial or in the 8 women with a pregnancy before and treatment with IL-2. Statistical comparison was performed using the chi-square test; p < 0.05 was considered significant.

Participants were first monitored and immunophenotyped during an untreated cycle. For the next cycle, they were administered 3 million international units (MIU) of IL-2 daily for 5 consecutive days, starting 10 days after the onset of menses (day 1) (**Figure 1A**). Of note, the 3MIU per-injection dose was chosen because it produces longer-lasting Tregs effects than lower, better-tolerated doses^33^. At this dose, a 5-day injection course maintains a Treg expansion for about a month ^19^. This regimen was timed so that IL-2 would be cleared by the potential time of embryo implantation, given a half-life of subcutaneaous IL-2 of about 4 hours and an embryo implantation at 6 to 10 days after ovulation, corresponding to days 20 to 24 of the standard menstrual cycle. If pregnancy was not achieved (β-HCG negative on day 29), a new treatment cycle was started, with a maximum of 5 cycles (**Figure 1A)**. Treatment cycles were planned as consecutive, but an interruption of up to two cycles was allowed. The dose of IL-2 could be decreased in case of side effects, but to no less than 1MIU/day.

### Immunomonitoring

The immune status of the FACIL-2 patients was assessed during the first untreated cycle and the subsequent IL-2_LD_ cycles. Blood samples were obtained for immunological tests immediately before the administration of IL-2_LD_. Sample were obtained daily from day 10 to day 14, and at day 17 and day 29 of each cycle. All the immunomonitoring procedures were performed as previously described and are described in the Supplementary methods^24,34–36^.

### Outcomes

The primary endpoint of the study was the increase in Treg count on day 14 of the first treatment cycle compared to baseline (day 10 of the untreated cycle). The secondary endpoints were the number of full-term pregnancies and safety assessed by the biological and clinical adverse events of the mother, fetus and newborn up to 6 weeks of age.

### Statistical analysis

Changes in Tregs between baseline and the other follow up timepoint were analyzed throughout the treatment period by non-parametric ANCOVA for repeated measurements, using baseline values as covariable (Conover method). Data are presented as mean ± SEM. Normal distribution of each set of variables was analysed by D’Agostino & Pearson omnibus normality test. Student’s t-test for normally distributed data and non-parametric Mann–Whitney were used to compare variables as appropriate. Paired data comparing follow-up time points to baseline value were analysed using Student’s t-test for normally distributed data and paired Wilcoxon signed-rank test for nonparametric. A p value < 0.05 was considered significant. Statistical analyses were performed using GraphPad Prism version 10.0 (GraphPad Software, San Diego, CA, USA).

### Compassionate use of IL-2_LD_ in REPL

During the same period, nine additional women who were unable or unwilling to participate in the FACIL-2 trial received the same IL-2_LD_ treatment on a compassionate use. They also had a history of at least 5 uREPLs before 14 weeks of gestation. These patients did not have the planned visits of the FACIL-2 study and had no biological follow up nor enforced safety follow up.

All patients treated at any AP-HP hospital (the body that oversees all public hospitals in the Paris region) are informed that data obtained during their care may be used for research purposes. Patients have the right to opt out if they wish. The nine additional patients mentioned did not opt out and are not included in a research protocol. The pregnancy outcome results that we report are from their care.

## RESULTS

### Overall study design and Patients

Fifteen women were enrolled in the FACIL-2 trial and completed a baseline (untreated) menstrual cycle followed by at least one IL-2_LD_ treatment cycle (**Figure 1A, 1B**). Of these, 9 received a second cycle, 3 a third one, 2 a fourth one and 1 a fifth one. The reason for discontinuation of the treatment was either pregnancy, patient preference or having missed more than 2 treated cycles. In parallel, 9 patients were treated on a compassionate basis (**Figure 1B**). Compassionate-use patients were not eligible for FACIL-2 due to exclusion criteria (e.g., age >40, autoimmune comorbidities) or declined trial participation due to logistical constraints. Of note, 2 of these patients were initially enrolled in FACIL-2 and were excluded from the trial after the first and the third cycle of IL-2_LD_, respectively, because they had a discontinuation of their treatment for more than 2 cycles. Of these 9 patients, 7 received a second cycle, 4 a third and 1 a fourth.

Inclusion criteria were consistent across both cohorts: women aged 18–40 with ≥5 uREPLs before 14 weeks of gestation. All these patients were treated with the same treatment modalities. Only the FACIL-2 patients were followed clinically, biologically and for safety. Baseline characteristics of both groups are shown in Table 1.

**Table 1.**
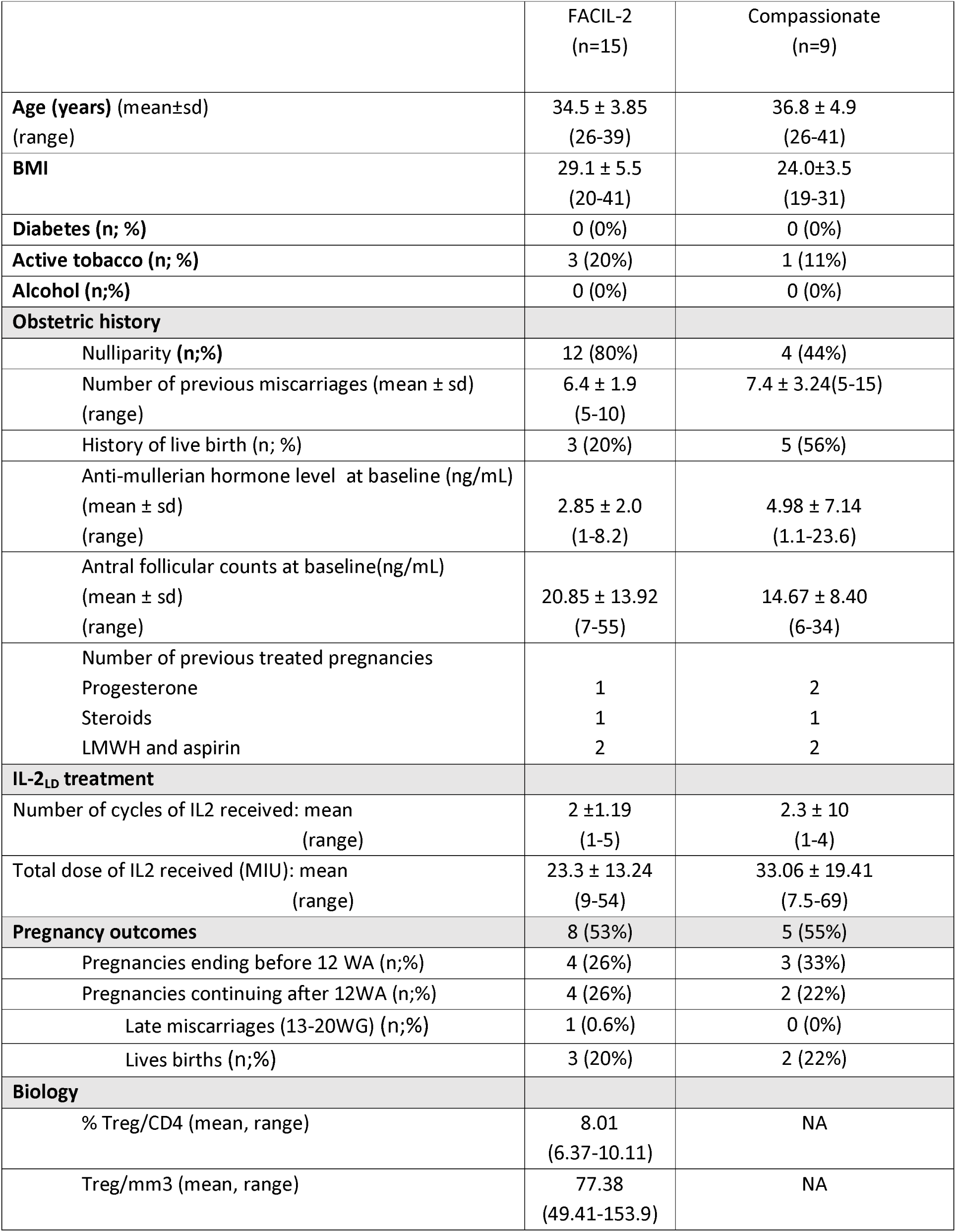
Demographic, clinical and biological characteristics.

### Treg response to IL-2_LD_

Before receiving their first IL-2 treatment cycle, FACIL-2 patients were monitored during an untreated cycle. Mean Tregs proportion remained relatively stable during this follow up, but with a slight but significant increase of Tregs percentage from 7.93±1.66 at day 10 to 8.49±1.72 at day 14 (p=0.016) (**Figure 2A**).

**Figure 2:**
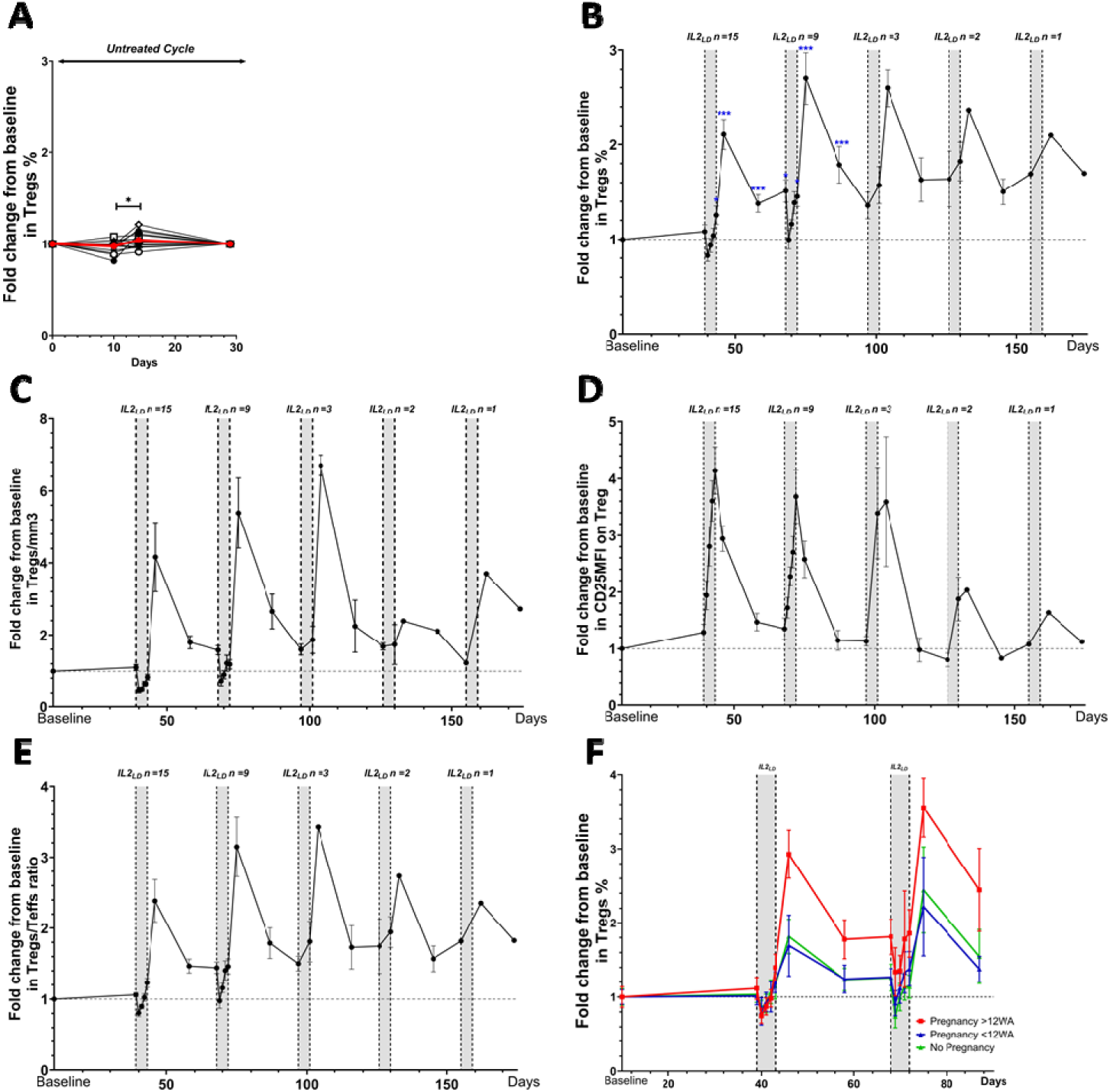
Changes in Regulatory (Treg) and Effector T cells (Teff) during IL-2_LD_ treatment. Treg cells were identified within the CD4⁺ T cell population as CD25^hiCD127^lo/-^Foxp3⁺ cells. (A–F) Changes from baseline are shown as either individual curves or as mean ± SEM: (A) Individual curves of Treg percentages among CD4⁺ T cells during the first treatment-free cycle, with the red line representing the mean across all patients; (B) Treg percentages among CD4⁺ T cells over the full follow-up period under IL-2_LD_ treatment; (C) Absolute counts of Tregs; (D) CD25 MFI in Tregs; (E) Tregs to Teffs ratio; (F) Comparison of Treg percentages during the first two treatment cycles stratified by pregnancy outcome: successful pregnancy (>12 WA, red, n = 4), early pregnancy loss (<12 weeks, blue, n = 4), and no pregnancy (green, n = 7).Statistical comparisons during follow-up were conducted using t-tests or Mann–Whitney tests, depending on data distribution (*p<0.05; **p<0.01; ***p<0.001).

In contrast, there was a marked Treg response to IL-2_LD_ during all treated cycle (**Figure 2B**).

During the first 5 days of the first treatment cycle, there was a small decrease of mean Treg proportions in the blood indicative of a Treg recirculation, as previously reported^23^. This was then followed by an increase that peaked at day-17 (8 days after treatment initiation), also as already noted^24,27^, with an overall 2.01±0.24 mean fold increase of mean Treg proportion (p<0.001). Thus, the primary criteria of the study has been reached. Of note, the increase persisted up to day-29 with a 1.33±0.24 fold increase. There was some heterogeneity in the response (**Figure S1**), as already reported^22,26^. These observations concerning the percentage of circulating Tregs were also true for their absolute number (**Figure 2C**). For the 2^nd^ cycle of treatment, the kinetic of the effect was similar, but both the peak value at day-17 (2.47±0.76 fold) and the persisting increase at day-29 (1.55±0.40 fold) were slightly higher than for the 1^st^cycle (**Figure 2B**). This was also true for the 3^rd^ cycle, and there were not enough patients going through the 4^th^ and 5^th^ cycle to draw conclusions (**Figure 2B**).

IL-2_LD_ also activated the expanded Tregs as shown by an increase of CD25 expression (**Figure 2D**)^33,24,26,36^. As there was no effect on effector T cells, the expansion of Tregs translated in a marked increase in the Treg/Teff ratio (**Figure 2E**).

### Clinical outcomes

In FACIL-2 participants, out of eight pregnancies, four were viable at 14 weeks, resulting in three live births and one late miscarriage at 20 weeks for a premature rupture of menbranes. Of note, this last patient had 19 previous pregnancy losses, always before 10 WG. In the compassionate-use group, five pregnancies occurred, yielding two additional live births. In the compassionate use group, five pregnancies resulted in two live births. All the born babies were in good health, with an APGAR score of ≥9 at 5 and 10 minutes.

There were no differences in the number of previous uREPL or in baseline Treg levels between the FACIL-2 patients who had viable and non-viable pregnancies (**Table 2**).

**Table 2:** clinical and biological outcomes in patients with succesfull pregnancy and pregnancy loss.

**Tableau 2:** clinical and biological outcomes in patients with succesfull pregnancy and pregnancy loss.
| Pregnancy | < 14 Week (N = 4) | ≥ 14 Week (N = 4) | p |
| --- | --- | --- | --- |
| Age at the pregnancy | 33.28±0.66 | 35.35±1.97 | ns |
| AFC | 31.0 ±18.02 | 11.5 ± 6.34 (n=2) | ns |
| AMH | 3.25±1.64 | 3.02±1.60 | ns |
| Number of previous early pregnancy loss | 7.25 | 6.75 | ns |
| Number of cycles of IL2 received: mean<br>(range) | 1.75±0.5<br>(1-2) | 1.75±0.5<br>(1-2) | ns |
| Total dose of IL2 received (MIU): mean<br>(range) | 17.13±9.06<br>(9-30) | 24.0 ±7.3<br>(15-30) | ns |
| Tregs at Baseline (mean ± sd; range) | 8.16±2.82<br>(5.07-9.91) | 7.41±1.19<br>(6.0-8.89) | ns |
| Tregs at entry in the first IL-2 cycle<br>(mean ± sd; range) | 8.43±1.58<br>(6.64-10.47) | 7.32±1.33<br>(5.61-8.67) | ns |
| Tregs fold increase at day-8 of the first and<br>second cycle (pooled values) | 1.81±0.51 | 2.84±0.51 | 0.03 |
| Tregs fold increase at day-29 of the first and<br>second cycle (pooled value) | 1.24±0.09 | 1.71±0.34 | 0.008 |

Notably, the levels of Treg activation and expansion appeared to be higher in patients who had successful pregnancies, with a Treg fold increase of 2.84 ± 0.51 vs. 1.83 ± 0.51 (p=0.03) at day-17. Moreover, the duration of the effect was also more pronouced with a Treg fold increase of 1.71 ± 0.34 vs 1.24 ± 0.09 (p= 0.008) on day-29, the end of the first cycle. (**Table 2 and Figure 2F**.

### Safety

There was a total number of 17 reported SAEs (**Table 3**). Of these 10 were deemed related to the IL-2 treatment. They were all observed at the dose of 3MIU of IL-2. There were two Hepatic cytolysis and two Flu-like syndromes with one associated with biological inflammation. Of note, six patients experienced cardiac symptoms including thoracic pain and/or EKG abnormalities (n=3 at 3MIU and n=3 at 1.5MIU). To our knowledge, such cardiac symptoms have never been reported in the many hundreds of patients with autoimmune diseases treated with IL-2, usually at dose ≤2MIU. These side effects led to the reduction of the IL-2 dose used to 1.5 or 1 MIU/injection. For one patient treatment was discontinued because of the EKG modification showing negative T waves.

**Table 3:** Safety.

|  | Nb Events (N° Patients)<br>[% administrations] | Nb Events 3MUI<br>(N° Patients) | Nb Events 1,5/1MUI<br>(N° Patients) |
| --- | --- | --- | --- |
| Nb of patients treated | 15 |  |  |
| Nb of injections administered | 138 (15) |  |  |
| **Serious Adverse events (SAE)** | **17 (10) [12,3%]** |  |  |
| SAE related to Proleukin® : | 10 (7) [7,2%] | 7 (6) | 3 (3) |
| Cardiac symptoms (thoracic pain and /or EKG abnormalities) | 6 (4) | 3 (3) | 3 (3) |
| Hepatic cytolysis | 2 (2) | 2 (2) | 0 |
| Influenza-like syndrome | 1 (1) | 1 (1) | 0 |
| Biological inflammatory syndrome associated with influenza-like syndrom | 1 (1) | 1 (1) | 0 |
| **Non Serious Adverse events (NSAE)** | **149 (15) [108%]** |  |  |
| NSAE related to Proleukin® : | 127 (15) [92%] | 105 (15) | 22 (4) |
| Grade 1 (73,2%) | 93 [67,4%] | 74 (14) | 19 (3) |
| Grade 2 (25,6%) | 33 [23,9%] | 30 (12) | 3 (2) |
| Grade 3* (7,8%) | 1* [0,7%] | 1 (1) | 0 |
| **Most common NSAE related to treatment** |  |  |  |
| Injection-site reactions | 45 (11) [32,6%] | 37 (11) | 8 (2) |
| Others AEs among which : | 82 (15) [59,4%] | 68 (15) | 14 (4) |
| Gastrointestinal symptoms | 21 (10) [15,2%] | 18 (9) | 3 (2) |
| Influenza-like syndrome | 19 (10) [13,8%] | 16 (10) | 3 (2) |
| Hypereosinophilia | 6 (6) [4,3%] | 4 (4) | 2 (2) |
| Mucocutaneous reactions | 6 (4) [4,3%] | 6 (4) | 0 |
| Biological inflammatory syndrome | 5 (4) [3,6%] | 5 (4) | 0 |
| Cardiac symptoms (thoracic pain, dyspnea, palpitations) | 4 (4) [2,9%] | 3 (3) | 1 (1) |
| Musculoskeletal pain | 4 (3) [2,9%] | 1 (1) | 3 (2) |
| Asthenia | 3 (2) [2,2%] | 2 (2) | 1 (1) |
| Headache | 3 (3) [2,2%] | 2 (2) | 1 (1) |
| Infection | 3 (2) [2,2%] | 3 (2) | 0 |
| Malaise | 2 (2) [1,4%] | 2 (2) | 0 |
| Fever | 1 (1) [0,7%] | 1 (1) | 0 |
| Hypokalemia* | 1* (1) [0,7%] | 1 (1) | 0 |
| Hepatic cytolysis | 1 (1) [0,7%] | 1 (1) | 0 |
| Loss of appetite | 1 (1) [0,7%] | 1 (1) | 0 |
| Hand paresthesia | 1 (1) [0,7%] | 1 (1) | 0 |
| Hot flushes | 1 (1) [0,7%] | 1 (1) | 0 |

\*hypokalemia corresponding to grade 3NSAE

There was a total 149 NSAE reported, with 127 deemed related to IL-2. For these later, 93 were Grade 1, 33 were Grade 2, and 1 was Grade 3. Of these, 105 were observed at the dose of 3MIU and 22 at the dose of 1.5 or 1 MIU (**Table 3**).

There were no adverse events for the fetus and there was no reported AEs during the baby follow-up, which occurred at 2, 3 and 6months.

## Discussion

Recurrent early pregnancy loss remains a major clinical challenge, particularly in cases where standard evaluations fail to identify a cause^37^. Here, we report the first human trial suggesting that low-dose IL-2 therapy can restore immune tolerance and improve pregnancy outcomes in women with unexplained REPL.

The rationale for this approach stems from accumulating evidence that immune dysregulation—specifically, Treg insuficiency—contributes to fetal rejection in uREPL. In animal models, Tregs are critical for establishing maternal-fetal tolerance. Ablation of Tregs in mice leads to increase rate of abortion^14,38,15,12^. Conversly, Treg activation and expansion by IL-2_LD_ has been shown to prevent fetal loss in the classical murine model of spontaneous abortion of DBA/2xCBA/J mating ^12,32,39^. In humans, a decrease in Tregs cells in the blood and endometrium and a reduced capacity to secrete IL-10 and TGF-β have been observed in women with uREPL ^3,5,9,10^. Thus, expanding and activating Tregs during early pregnancy appears as a valid therapeutic target for uREPL. Moreover, any positive outcome of such treatment will contribute to establish Treg insuficiency as a major determinant of uREPL.

Our study demonstrates for that IL-2_LD_ can safely expand and activate Tregs during the peri-implantation window. The primary efficacy criteria of the study – the increase of Treg proportion at day-14 that peak at day-17 of the first cycle – was successfully met. Indeed, 5 consecutive days of IL-2 at 3MIU led to robust and significant Treg expansion, with an average 2-fold increase by day 8 after treatment initiation. Such increase, as well as the initial decrease of Treg values during the first 2-3 days, is in agreement with the effects of similar dose of IL-2_LD_ in patients with other conditions and in healthy volunteers^19,22,33,26,24,40,28,27,29,30^. Thus, Tregs from pateints with uREPL respond normaly to IL-2_LD_.

In this proof of concept study, we chose to treat patients for only 5 days starting at day 10 of the menstrual cycle. Indeed, as IL-2 has initially been approved for treating advanced cancers, it has not been subjected to a formal genotoxicity study. Although genotoxicity for a natural molecule is very unlikely, it appeared preferable to not expose embryos to IL-2. As the half-life of IL-2 is of about 4 hours, and the implantation of the embryo at day 20 to 24, the administered IL-2 would have been washed out before implantation. However, the duration of the biologic effect on Tregs being much longer that its half life, the Treg stimulation was expected to cover about 2 to 4 weeks after the last injection. Although we would have liked to cover the entire first trimester of pregnancy, this schedule of administration appeared as a safe one for a first administration in this clinical setting. We indeed observed that during the first cycle, the effects on Tregs lasted for at least 4 weeks, with increased values at day-29 and at day-10 of the next cycle. This expansion of Tregs was accompanied by their activation as evaluated by the level of CD25 epression, a well known marker of Treg activation. The activation level increased everyday during treatment, and rapidely vanished as already reported. Altogether, patient with uREPL respond to IL-2_LD_ as do all others^19–22,33,24,26,40,27,29,30,25^.

Some patients had reduction of their IL-2 dosage to 1.5 or 1MIU/injection due to tolerance issues (see below). This mostly occurred at different time points of treatment, although mostly after the first cycle of treatment. The heterogeneity of the IL-2 dose adjustment (dose and time) does not allow to draw conclusion such as to the compared effects of different IL-2 dosage in the context of uREPL treatment.

In previous studies, we evaluated the safety and efficacy of Treg activation by dose of IL-2 up to 3MIU. Although the effects of 3MIU were stronger and lasted longer ^19,22,33^, we therafter used IL-2 mostly at dose up to 2MIU because of a better tolerance. Indeed, IL-2 has a favorable safety profile at all dose up to 3MIU/injection, with mostly mild AEs, but there are still more AEs at the 3MIU dosage^22,41^. Here we started at 3MIU to maximise the duration of the effect, which is does dependant, and had to decrease the dosage in 7 patients. While most adverse events were mild and expected ^41,42^, transient cardiac symptoms in a subset of patients receiving 3 MIU/day were unanticipated and not observed in previous IL-2_LD_ trials for autoimmune diseases. The etiology remains unclear but prompted a dose adjustment to 1.5–1 MIU/day without apparent loss of efficacy on Treg effects.

This rate of successful pregnancies appears higher than expected in such a severely affected population20, especially considering that only around 50% of uREPL are thought to have an immunological origin. The 15 women enrolled in FACIL-2 collectively experienced 99 previous pregnancies, of which only three ended in live birth, providing an estimate of the baseline rate of successful pregnancies in this population. In contrast, of the eight women who became pregnant during the trial, three achieved live birth and one experienced pregnancy failure for mechanical reasons beyond 14 WG. This rate of successful pregnancies under IL-2LD is significantly higher than their historical rate (p < 0.0008) and than the overall baseline rate in the FACIL-2 cohort (p < 0.0001) (**Figure 1C**). Notably, successful pregnancies in FACIL-2 were associated with significantly greater Treg expansion (2.87-fold vs 1.64-fold at day-17, p = 0.03; 1.71-fold vs 1.24-fold at day-29, p = 0.008) (Fig.2D). Collectively, these findings indicate that IL-2LD improves pregnancy outcomes in uREPL and that this benefit is linked to effective Treg stimulation.

When taking into account the compassionate use of IL-2 in a similar population, the pregnancy success rate is overall of 46%. As it is believed that about half of the uREPL do not have a immunological origin, this is a remarquable finding. This is particularly encouraging given that the intervention was both brief and not continued during gestation, a precautionary strategy that now warrants reconsideration given the therapy’s safety profile. The observation that the higher Treg expansion in successful pregnancy was significant up to day-29 support that longer treatment duration could be beneficial. Longer treatment duration will also allow using lower doses of IL-2 (1-1.5MIU) that have a better safety profile. Whether ongoing IL-2_LD_ treatment through the first trimester might further improve outcomes is a compelling hypothesis for future trials.

This study has several limitations. First, the open-label design and lack of a control group preclude definitive conclusions on efficacy. Second, the small sample size limits generalizability. Third, Treg profiling was not performed in the compassionate-use group. Finally, while IL-2_LD_ likely addresses an immune cause of miscarriage, not all uREPL cases are immunological, which may dilute apparent efficacy.

Despite these limitations, our results establish proof of principle that immunotherapy with IL-2_LD_ can restore maternal-fetal tolerance and lead to successful pregnancies in a high-risk population. A randomized, placebo-controlled trial of IL-2_LD_ administration extending into early pregnancy is now warranted to confirm efficacy and identify predictors of response.

## Data Availability

All data produced in the present study are available upon request to the authors

## Acknowledgement

We thank the patients for their participation in the trial and the personnel of the Pitié-Salpêtrière Clinical Investigation Center for Biotherapies (CIC-BTi), Michèle Barbié, Natalie Féry, Catherine Ferrapie, Nadia Graffin, Aurélie Marc, for their excellent technical assistance.

## Authors’ contribution to the current study

DK conceived and supervised the study.

AM, EV, MR and DK designed the clinical study.

NA, CM, CR and RL performed the clinical follow-up of patients under the supervision of AM.

MR supervised the immunomonitoring.

AM analysed the clinical results.

MR and DK analysed the biological results.

MR and DK wrote the first draft of the manuscript that was edited by all authors.

