## Supplemental appendix for "Low-dose interleukin-2 for recurrent early pregnancy loss: a proof-of-concept study"

**SUPPLEMENTARY APPENDIX**

**TABLE OF CONTENT**

- Table S1: Inclusion/Exclusion criteria
- Figure S1: Biological response (Individual Treg response)
- Supplementary Methods

**Table S1:** Inclusion/Exclusion criteria

| **PATIENTS**  **Main inclusion criteria** | - Female aged [18-40] years - Woman with at least 5 consecutive early miscarriages less than 14 weeks of amenorrhea and unexplained after the usual check-up; - Volunteer to participate in the trial and having given written consent after appropriate information. |
| --- | --- |
| **Main non-inclusion criteria** | - Uterine or pelvic abnormality: uterine malformation, intracavitary fibroid, synechiae, polyp, hydrosalpinx; - Balanced translocations in both spouses; - Diabetes type I or II; - Sickle cell disease; - Contraindication to pregnancy; - Constitutional or acquired thrombophilia (protein deficit C, S, ATIII, homozygous factor V or II deficiency, antiphospholipid syndrome, antithyroid antibodies positive, celiac disease, hyperhomocysteinemia); - Decreased ovarian reserve (AMH <1 ng/ml); AFC < 4 - Significant spermogram abnormalities and DNA fragmented more than 30% - Active HIV or HCV infection; - Main known contraindications to treatment with IL-2: - Hypersensitivity to the active substance or to any of the excipients; - Signs of progressive infection requiring antibiotic therapy; - History of organ allograft; - Pre-existing autoimmune disease; - Leukocytes <4000 / mm3; platelets <100,000 / mm3; hematocrit <30%; - hepatic or renal insufficiency; - depression; - significant history or existence of a serious heart disease (in doubtful cases, perform a stress test); - patients with autoimmune disease; - patients with an infection (septicemia, bacterial endocarditis, septic thrombophlebitis, peritonitis and pneumonia); - pregnancy; - Treatment with immunomodulators, immunosuppressants (class L04A of the ATC classification), in particular systemic corticosteroids, as well as aspirin and low molecular weight heparin; - No affiliation to a social security; - Person who has already been included in this study or in another at the same time; - Major incapacitated patient (tutorship / curatorship); - Patient with an allergy to taking IL2-fd; - Participants who would present professional risk factors (eg ionizing exposure); |

**Figure S1:** **Individual Treg response all along the follow up from the first cycle to the fifth cycle ( C1 n=15; C2 n=9 ; C3 n=3 ; C4 n=2; C5 n=1)**

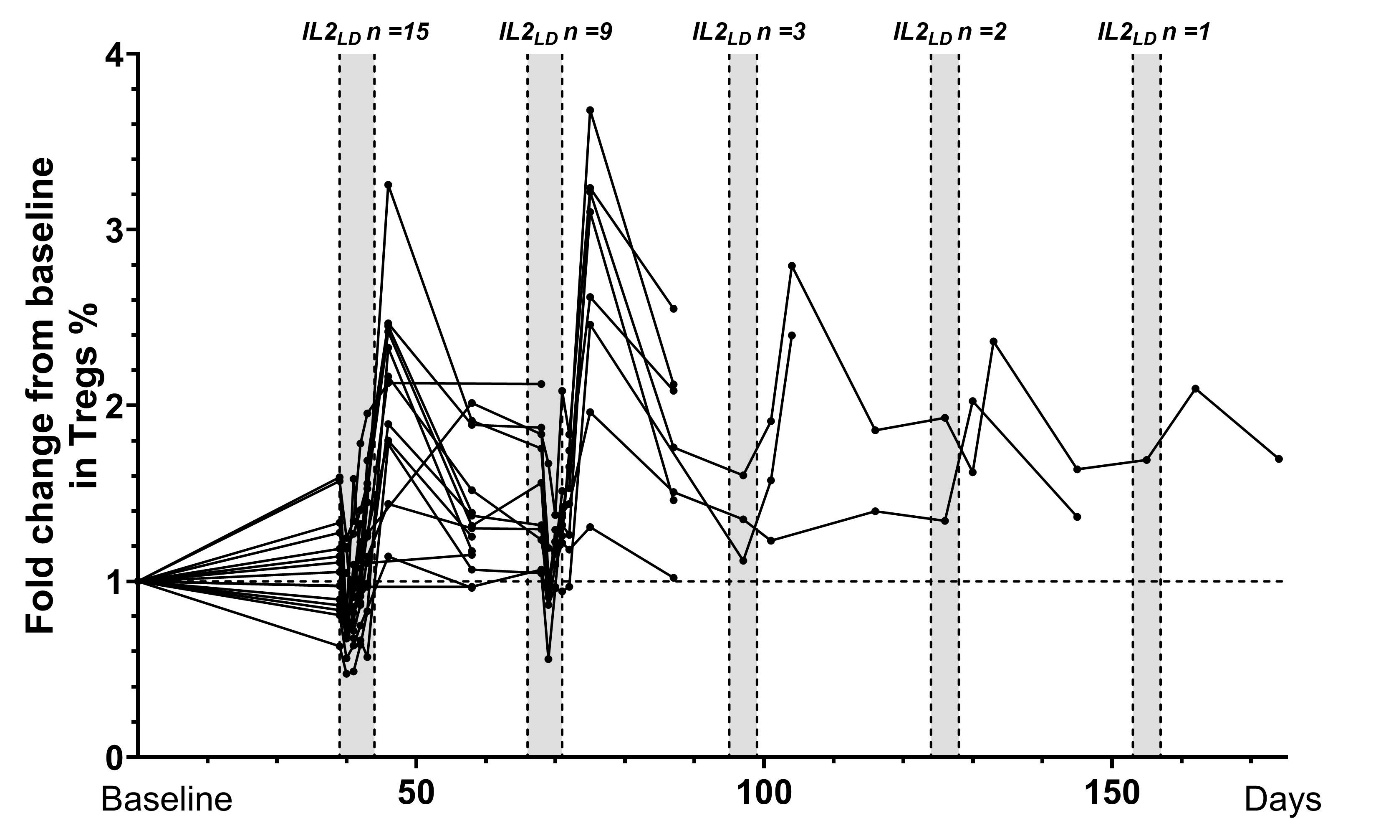

**Supplementary methods**

***Flow cytometry***

Blood samples were collected in EDTA tubes according to the planned protocol: absolute numbers of lymphocyte subsets and Tregs were monitored at each patient visit: day-10, day 17 and day 29 of the first of the cycle without treatment, for the other cycles at day 10 before treatment (baseline) at day 14 just before the fifth infusion of IL-2LD, day 17 and day 29 (at the end of the cycle).

Blood subsets (CD3^+^, CD4^+^, CD8^+^ T lymphocytes, CD19^+^ B lymphocytes and CD3^-^CD56^+^ NK cells) counts (cells/μl) were established from fresh blood samples using CYTO-STAT tetraCHROME kits with Flowcount fluorescents beads and tetra CXP software with a FC500 cytometer (Beckman Coulter) according to manufacturer’s instructions. Foxp3 labeling was performed with custom designed dry tubes (duraclone from Beckman Coulter) containing CD3-FITC, CD25-PE, CD127-PE-Cy7, Foxp3-AF647, CD8-KRO, CD8-PB antibodies as described already described^1^. Total blood (60 µL) sampled with anticoagulant was mixed with 6 µL of Perfix-NC R1 buffer, vortexed immediately for 2-3 seconds and incubated for 15 min at room temperature in the dark. Perfix-NC R2 buffer (360 µL) was added and, after homogenization, a 360 µL aliquot was transferred to a Duraclone tube. After vortexing for 10 seconds, tubes were incubated for 60 min at room temperature in the dark. PBS 1X (3 mL) was added to the tubes, incubated for 5 min at room temperature in the dark before centrifugation for 6 min at 250g. The supernatant was removed to leave the pellet dried and the cells were resuspended in 3 mL of 1X Perfix-NC R3 buffer prior to another 6-min centrifugation at 250g. The pellet was dried and resuspended in 300 µL of 1X R3 buffer. Tubes were protected from light and stored at 4°C until the acquisition on a cytometer within the next 24 h. Acquisition was performed on a Navios cytometer (Beckman Coulter) maintained daily according to the manufacturer’s recommendations with Flow Check Pro and Flow Set Pro fluorospheres. Acquisitions were performed, respectively, using Navios software, and all analyses were done with Kaluza 1.3 software (Beckman Coulter)^1^.
